# Integration of GWAS and multi-omic QTLs identifies uncharacterized COVID-19 gene-biotype and phenotype associations

**DOI:** 10.1101/2024.09.05.24313137

**Authors:** Meritxell Oliva, Emily King, Reza Hammond, John S. Lee, Bridget Riley-Gillis, Justyna Resztak, Jacob Degner

## Abstract

To better understand COVID-19 pathobiology and to prioritize treatment targets, we sought to identify human genes influencing genetically driven disease risk and severity, and to identify additional organismal-level phenotypes impacted by pleiotropic COVID-19-associated genomic loci. To this end, we performed ancestry-aware, trans-layer, multi-omic analyses by integrating recent COVID-19 Host Genetics Initiative genome-wide association (GWAS) data from six ancestry endpoints - African, Amerindian, South Asian, East Asian, European and meta-ancestry - with quantitative trait loci (QTL) and GWAS endpoints by colocalization analyses. We identified colocalizations for 47 COVID-19 loci with 307 GWAS trait endpoints and observed a highly variable (1-435 endpoint colocalizations) degree of pleiotropy per COVID-19 locus but a high representation of pulmonary traits. For those, directionality of effect mapped to COVID-19 pathological alleles pinpoints to systematic protective effects for COPD, detrimental effects for lung adenocarcinoma, and locus-dependent effects for IPF. Among 64 QTL-COVID-19 colocalized loci, we identified associations with most reported (47/53) and half of unreported (19/38) COVID-19-associated loci, including 9 loci identified in non-European cohorts. We generated colocalization evidence metrics and visualization tools, and integrated pulmonary-specific QTL signal, to aid the identification of putative causal genes and pulmonary cells. For example, among likely causal genes not previously linked to COVID-19, we identified desmoplakin-driven IPF-shared genetic perturbations in alveolar cells. Altogether, we provide insights into COVID-19 biology by identifying molecular and phenotype links to the genetic architecture of COVID-19 risk and severity phenotypes; further characterizing previously reported loci and providing novel insights for uncharacterized loci.

## Introduction

Since the beginning of 2020 till mid 2023, SARS-CoV-2 infection and its disease, COVID-19, have caused the largest contemporary pandemic to date, severing the life of more than 7 million people, completely disrupting society lifestyle dynamics and causing a world-wide financial crisis. On May 5, 2023, the World Health Organization (WHO) declared COVID-19 to be no longer a public health crisis worldwide, as vaccination efforts proved successful in neutralizing existent variants at time ^1^. However, due to continuous evolution and adaptation, SARS-CoV-2 new variants keep arising ^2,3^ . It is possible that forthcoming variants may escape neutralization of current vaccine approaches, as Omicron variant did for first generation vaccines ^2^. Even if up to date with vaccination, certain populations, like the immunocompromised, are disproportionally affected by the disease. Moreover, infection by SARS-CoV-2 can result in Long COVID, a condition characterized by symptoms of fatigue and pulmonary and cognitive dysfunction ^4^. Recent studies identified immunologic signatures linked to Long COVID symptoms ^5,6^, but the biological mechanisms that contribute to the development of the disease remain to be clarified. There is, thus, a need to keep further characterizing the host factors that contribute to COVID-19 etiology.

During early stages of the pandemic, it was made evident that COVID-19 presentation was highly variable and influenced by multiple demographic, environmental, and biological factors such as age, sex, race, presence of comorbidities and differential immune response ^7^. As observed for other viral infections, COVID-19 is influenced by host genetics ^8^. Three months after the pandemic was declared by WHO (March 11th, 2020) pioneer studies reported 9q34(ABO) and 3p21.31(SLC6A20) as genetic susceptibility loci for the development of respiratory failure in COVID-19. Global efforts devoted to unveiling COVID-19 genetics were put in place, notably spearheaded by the Host Genetic Initiative (HGI) ^9^. To date, as per GWAS catalog ^10^ entries (July 2024), more than 30 Genome-wide association studies (GWASs) have been conducted worldwide, mostly in European ancestry cohorts, identifying dozens of genomic loci linked COVID-19 phenotypes, mainly susceptibility and severity of COVID-19.

Comprehensive characterization of the shared genetics among COVID-19 and other complex traits has provided insights into the pathobiology of the disease and can inform repurposing of existing drugs. Phenome-wide approaches have identified COVID-19 links with body mass index, smoking, diabetes and ischemic stroke, among others, and causal relationships between COVID-19 and some of these traits have been predicted. For instance, genetic liability to increased BMI and bad smoking habits is reported to be causally linked to COVID-19 severity; whereas genetically determined better kidney and lung function might be protective ^11,12^. Such findings have implications for risk stratification in patients with COVID-19 and the prevention of its severe outcomes.

To identify molecular implications of GWAS signal, a straightforward approach is to consider genes based on their proximity to the risk loci. This approach is not truly accurate ^13^ and is outperformed by integration of quantitative trait loci (QTLs), which enables higher accuracy and can be informative of the causal context, e.g. cell type of origin ^14^. Several studies have utilized QTL-based approaches to prioritize putative causal COVID-19 genes ^15–18^. Collectively, they have provided supporting evidence for genes involved with mucociliary clearance, viral-entry and mucosal immunity as linked to of SARS-CoV-2 infection, and genes involved in antiviral immune response, leukocyte trafficking and lung injury as linked to severe disease^19^. However, most published approaches are derived from only expression QTLs (eQTLs) and restricted to a relatively scarce set of biotypes, mainly bulk-tissue lung, blood and immune cells. Importantly, the effects of common genetic variants on molecular phenotypes can be specific to a cell-type or other context, or observed in a particular molecular phenotype other than gene expression ^20–22^. Thus, integration of a wide range of multi-omic, multi-context QTL sources, including lung QTLs at a single cell resolution, can maximize the expectation to identify COVID-19 putative molecular mediators.

Here, we sought to identify human genes influencing genetically driven disease risk and severity, and to identify additional organismal-level phenotypes impacted by pleiotropic COVID-19-associated genomic loci. To this end, we performed ancestry-aware, trans-layer, multi-omic analyses by integrating recent COVID-19 Host Genetics Initiative genome-wide association (GWAS) data from six ancestry endpoints - African, Amerindian, South Asian, East Asian, European and meta-ancestry – with expression, splicing, protein and DNA methylation QTLs, and with GWAS endpoints, by systematic colocalization analyses.

## Results

### Overview and characterization of COVID-19 linked genomic loci

We analyzed ancestry-stratified and multi-ancestry GWAS data provided by the COVID-19 Human Genetic Initiative v.7 ^11,23^ and identified 91 loci putatively associated with COVID-19, defined as genomic loci with suggestive (P<5-07) association with COVID-19 disease susceptibility and/or severity phenotypes (Supplementary Table 1). For 38 loci, no association with COVID-19 phenotypes has previously been reported (Methods), but only 8% (3/38) are genome-wide significant (P<5-08). Out of the 91 COVID-19 loci, 28% were exclusively identified in an ancestry-stratified GWAS endpoint; 68% of those (17/25) in non-European cohorts. We observe, as arguably expected, that ancestry-specific loci tend to correspond to low-frequency variants with relatively higher frequency in the discovery cohort, but with consistent direction of effect across GWAS endpoints (Fig. 1).

**Fig. 1.**
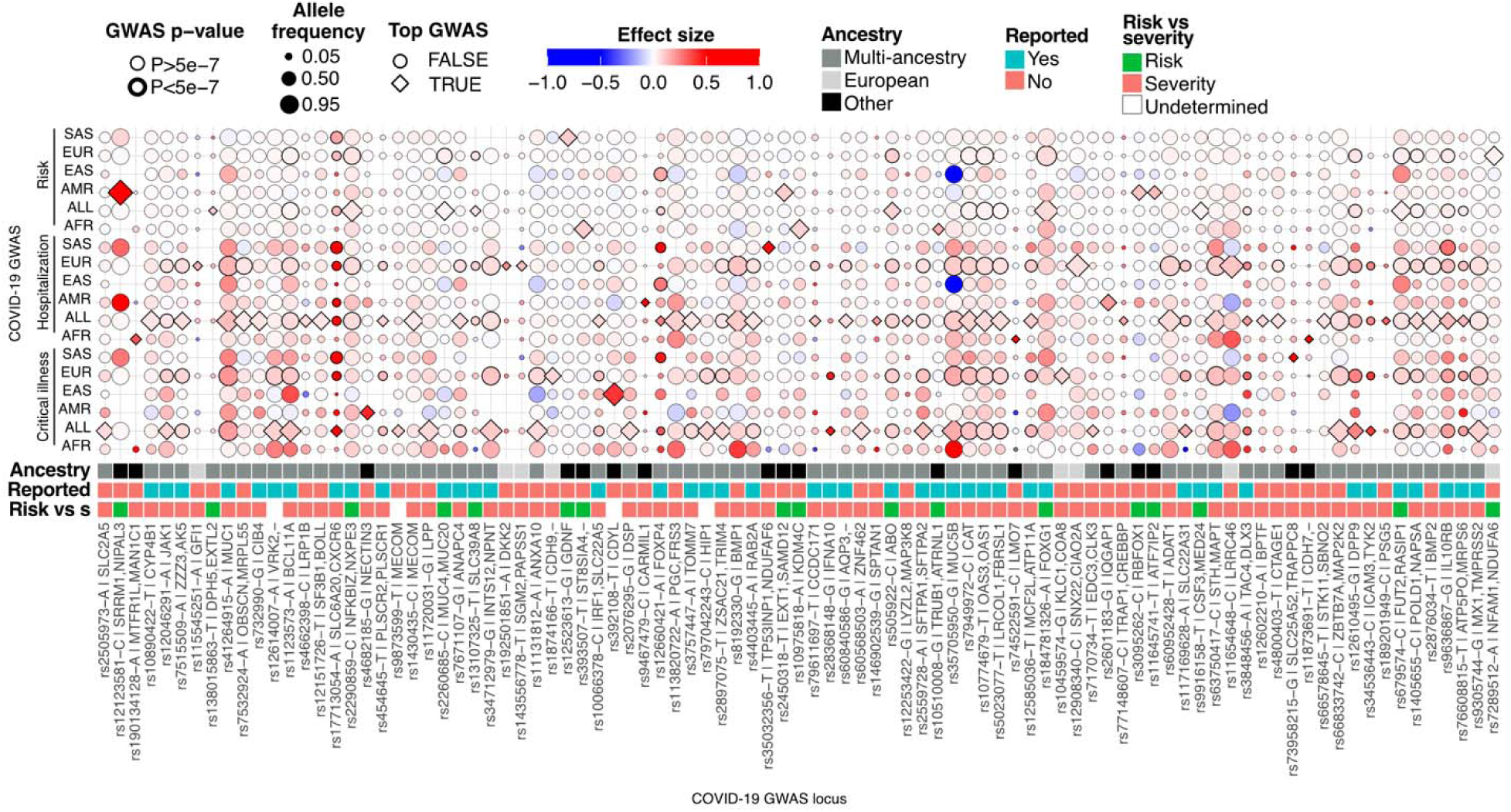
Genome-wide associations for SARS-CoV-2 risk and COVID-19 severity across ancestries. Suggestive (P<5-07) genomic loci (x axis) associations with COVID-19 severity (Hospitalization, Critical illness) and SARS- CoV-2 susceptibility (Risk) phenotypes across ancestry (ALL=Multi-ancestry, AFR=African, AMR=Admixed American, EAS=Southeast Asian, EUR=European, SAS=South Asian) endpoints (y axis). COVID-19 loci are labeled by lead variant, risk/severity (pathological) allele, corresponding closest protein-coding gene, and Open Targets V2G gene (if different). Top GWAS is indicated by a rhomb and corresponds to the GWAS with smallest lead variant p-value per locus. Loci are annotated by top GWAS ancestry, ‘Reported’ for literature reported-status, and risk versus severity score (Methods, Supplementary Table 1). Circle size represents the allele frequency of the pathological effect allele. Circle color and boldness represent GWAS effect size and significance (P_GWAS_<5-07), respectively.

For 46 loci, corresponding lead variant had suggestive (P<5-07) association across multiple GWAS endpoints. For those cases, to avoid testing correlated GWAS signal per locus, only the GWAS with the most significant signal per locus was considered for subsequent analysis (Fig. 1, Methods). To identify biological signatures associated to different COVID-19 features in downstream analyses, we employed a two-class Bayesian model ^12^ and determined that most classified (80%, 70/87) loci are likely to influence severity of the disease (Risk vs severity score<0.2) and a substantial minority (20%, 17/87) are more likely to impact susceptibility to SARS-CoV-2 infection (Risk vs severity score>0.8) (Fig. 1). This proportion matches previous observations with stricter threshold (score<0.01 or >0.99) ^11^.

Previously reported COVID-19 GWAS genes and loci are highly represented in viral entry, entry defense in airway mucus, type I interferon response and upkeep of healthy lung tissue. Here, among previously unreported COVID-19 loci, we identified additional instances to such processes by considering putatively implicated genes by COVID-19 GWAS lead variants given closest gene, VEP^24^ and V2G^25^ variant-to-gene approaches (Methods, Supplementary Table 1, Fig. 1). For instance, we identified rs2076295 as predicted to impact *DSP*, which encodes a desmoplakin linked to pulmonary diseases and is thought to maintain airway epithelial integrity ^26^. We observed rs8192330 overlap with *SFTPC* isoforms, which regulate alveolar surface tension and were found to be significantly decreased in COVID-19 patients with higher viral load ^27^. Unreported loci also pinpoint genes and mechanisms for which the COVID-19 link is less characterized, but that are arguably good candidates for playing a role in the disease. For instance, rs60568503 is located in an intronic region of the long non-coding RNA gene *LINC01505*, which is predicted to interact with SARS-CoV-2 spike mRNA ^28^. The variant rs190134128, only common (MAF>0.01) in African cohort, is located in an intronic region of *SELENON*, which encodes selenoprotein N; selenium levels have been correlated with COVID-19 survival and selenoproteins levels are altered by SARS-CoV-2 infection ^29^. The variant rs60952428 is predicted to impact *ADAT1*, which encodes for a deaminase responsible for the deamination of adenosine 37 to inosine in eukaryotic tRNA; A-to-I RNA editing is increased in COVID-19 infected individuals ^30^. The variant rs2505973 is predicted to impact *SLC2A5,* which encodes an enzyme involved in fructose uptake, is differentially expressed in SARS-CoV-2 infected cells and is hypothesized to play a role in upkeeping their energy needs ^31^. We also identify loci associated to proteins that are part of the same complex or that physically interact. For instance, variants rs9873599 and rs1430435, are predicted to impact *MECOM*, which can interact with *CREBBP*, associated to variant rs77148607. Both genes are involved in cell cycle and differentiation and cancerous processes ^32,33^. The variants rs7289512 and rs35032356 are predicted to impact *NDUFA6* and *NDUFAF6*, which are assembly- and structure-associated proteins of the mitochondrial respiratory complex I, respectively. Mitochondrial dysfunction has been associated to COVID-19 pathobiology ^34,35^. Overall, characterized genomic loci potentially associated to COVID-19 pinpoint known and novel or less characterized COVID-19 putatively causal genes and mechanisms.

### Identification and characterization of pleiotropic COVID-19 loci

Comprehensive characterization of shared genetics between diseases can provide valuable insights into their pathobiology ^36^. Genome-wide cross-trait studies have found evidence for shared genetic architecture between COVID-19 and BMI, diabetes, smoking habits, cardiovascular and pulmonary traits, among others ^11^, but availability of GWAS summary statistics has historically been a limiting factor. To characterize the shared genetic basis of COVID-19 with a broad spectrum of trait endpoints, we performed COVID-19 GWAS colocalization with other GWAS traits. We used *POEMcoloc* ^37^, an imputation-colocalization approach that infers shared causal genetics of two traits from as little as summary statistics from a single lead variant. We integrated and colocalized COVID-19 GWAS signal with GWAS summary statistics from the entire GWAS Catalog (February 2023) ^10^, comprising 89 COVID-19 loci overlapping with 13,750 loci from 1,053 GWAS trait endpoints (Methods).

In total, we identified significant (PP4>0.75) colocalizations derived from 307 GWAS trait endpoints and corresponding to 47/89 (53%) of the COVID-19 loci, 19 of which were not previously reported as linked to COVID-19 phenotypes (Supplementary Table 2, Fig. 2a). We observed a highly variable degree of pleiotropy per COVID-19 locus, and a tendency for COVID-19 susceptibility loci to be more pleiotropic than severity loci, as we observed a correlation between the number of colocalized traits per locus and the corresponding risk versus severity score (Spearman rho=0.56, P<1e-04). Two notable examples include the susceptibility-linked loci *SLC39A8* and *FUT2*, which colocalize with more than 150 trait endpoints across multiple trait classes (Fig. 2a, left panel).

**Fig. 2.**
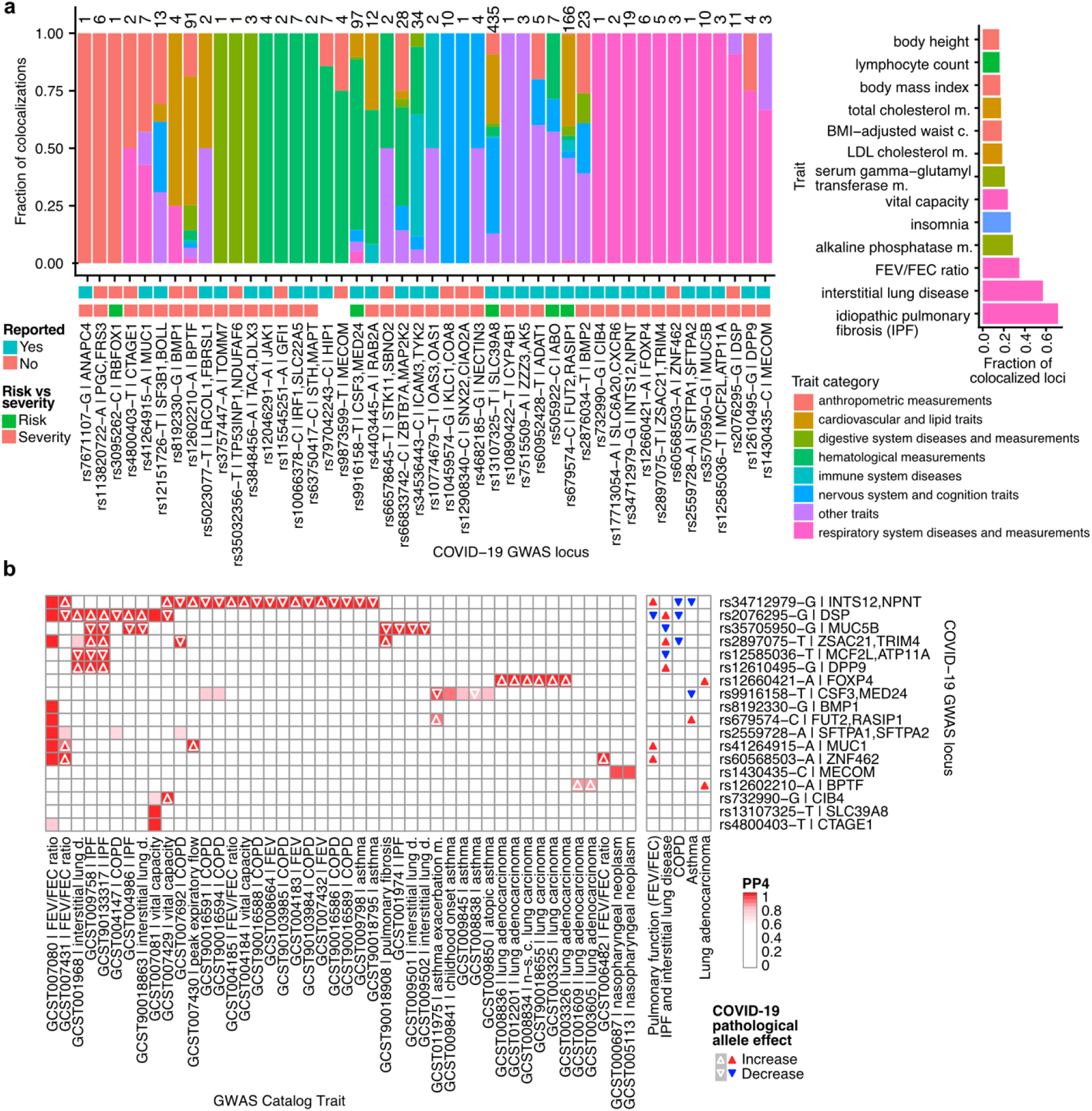
Pleiotropic COVID-19 linked loci. **a**, Leftmost bar plot represents the fraction of colocalizations (y axis) corresponding to each COVID-19 pleiotropic locus (x axis) stratified by broad category of GWAS Catalog trait. Total number of colocalizations per locus is displayed on top of corresponding bar. COVID-19 loci are labeled as in figure 1 and annotated by ‘Reported’ for literature reported-status, and risk versus severity score (Methods, Supplementary Table 1). The rightmost bar plot represents the fraction of GWAS Catalog loci that colocalize with COVID-19 (x axis) per trait narrow category (y axis). For a given category, the fraction is the number of overlapping GWAS Catalog loci mapped to the trait category for which a significant (PP4>0.75) colocalization is observed. Details of the colocalization approach and trait category mapping are described in Methods. **b**, Leftmost heatmap represents *POEMColoc* probability of colocalization (PP4) per COVID-19 locus (y axis) and respiratory trait endpoint (x axis) for loci that colocalize with at least one respiratory trait. COVID-19 pathological allele effect on pulmonary trait is depicted with an upward (increase) or downward (decrease) triangle, for cases with available data. Rightmost heatmap represents consensus pathological allele effect on trait derived from aggregated evidence across endpoints.

Colocalizations with COVID-19 are prevalent among respiratory system related traits, cardiovascular and lipid related traits, anthropomorphic measurements and hematological measurements, which reflects, in part, their higher overall prevalence in the GWAS Catalog. To identify highly represented trait classes, we considered the fraction of significant COVID-19 colocalizations within overlapping GWAS loci (Methods). We observed that overlapping associations for pulmonary traits were most frequently colocalized (fraction>0.3), with idiopathic pulmonary fibrosis (IPF), interstitial lung disease (ILD) and pulmonary function measurements being the top represented traits (Fig. 2a, right panel). These findings are in line with what has been observed ^11^, and highlights that the genetic predisposition to COVID-19 risk or severity shares more genetic basis with pulmonary phenotypes compared to other traits and that there is a systemic dependence on genetic factors influencing pulmonary biology.

To characterize the individual contribution of each COVID-19 locus on associated pulmonary traits, we analyzed the impact of COVID-19 pathological alleles on the direction of effect per trait, imputing it from variants in linkage disequilibrium when not directly available (Methods, Fig. 2b). We observed that COVID-19 pathological alleles can be either protective or detrimental for other traits, and oftentimes per-locus opposite effects are observed within traits. We link the rs12660421-A in the *FOXP4* locus to increased lung adenocarcinoma risk and note that this allele has recently been reported to be associated to long COVID-19 ^38^. At each of three colocalizing loci, COVID-19 pathological alleles convey protective effects for chronic obstructive pulmonary disease (COPD), despite ILDs and COPD being a risk factor for COVID- 19 ^39^. The COVID-19 pathological alleles rs35705950-G and rs12585036−T in the *MUC5B* and *MCF2L|ATP11A* loci, respectively, have been found here, and previously, to be protective for ILDs ^40,41^. On the other hand, observed IPF risk alleles include the COVID-19 pathological alleles rs2897075−T | *ZSAC21|TRIM4*, rs12610495−G|*DPP9* and rs2076295-G|*DSP*. These loci are known to contribute to IPF genetic architecture ^42^, but the *DSP* locus has not been previously associated with COVID-19. Observed effects on pulmonary function metrics were locus-dependent as well. For example, rs41264915-A|*MUC1*, rs34712979-G|*INTS12|NPNT* and rs60568503−A|*ZNF462* were linked to increased pulmonary function (FEV/FEC ratio), but rs2076295-G|*DSP* showed the opposite effect. Altogether, our results contribute to the understanding of COVID-19 genetic mechanisms shared with other traits, particularly with pulmonary pathologies.

### Impact of COVID-19 GWAS loci on molecular phenotypes

To infer molecular consequences of COVID-19 GWAS variants, we integrated by colocalization a comprehensive set of 284 *cis* QTL maps derived from different molecular phenotypes, 149 bulk-tissue and 135 cell-isolated sources. This set comprises maps from gene expression or splice phenotypes (e/sQTLs, 94%), DNA methylation (mQTLs, 4%) and protein (pQTLs, 2%) abundance, including in disease-relevant biotype and contexts like lung, plasma, blood and immune cells (Methods, Supplementary Table 3). Across all COVID-19 associated genomic loci, QTL maps and molecular phenotypes, we identified 2,511 colocalizations (PP4>0.75, Supplementary Table 4) across 64 loci. While 83% of colocalized loci implicate at least one eQTL endpoint, considering loci with QTL colocalization(s) identified in other molecular phenotypes, 11/62 lack eQTL associations. Molecular QTL-colocalized loci involved most reported (47/53) and half of unreported (19/38) COVID-19-associated loci, including 9 loci identified in a non-European ancestry (Fig. 3a). Among these non-European loci, we observed the COVID-19 risk allele rs10975818-A, identified in the African-ancestry cohort, associated with higher *KDM4C* expression in T-cells, and the rs2601183-G COVID-19 severity allele, identified in the Amerindian cohort, associated with higher CpG-site methylation levels in *ZNF774* 3’ exon in lung (Fig. 3b). Higher expression of *KDM4C* has been linked to lung cancer ^43^, and *ZNF774* expression is enhanced in glandular epithelial cells and predicted to be associated with mucin production ^44^. By analyzing ancestry-stratified GWAS signal and multi-omic QTLs, we revealed molecular links to previously uncharacterized COVID-19 loci.

**Fig. 3.**
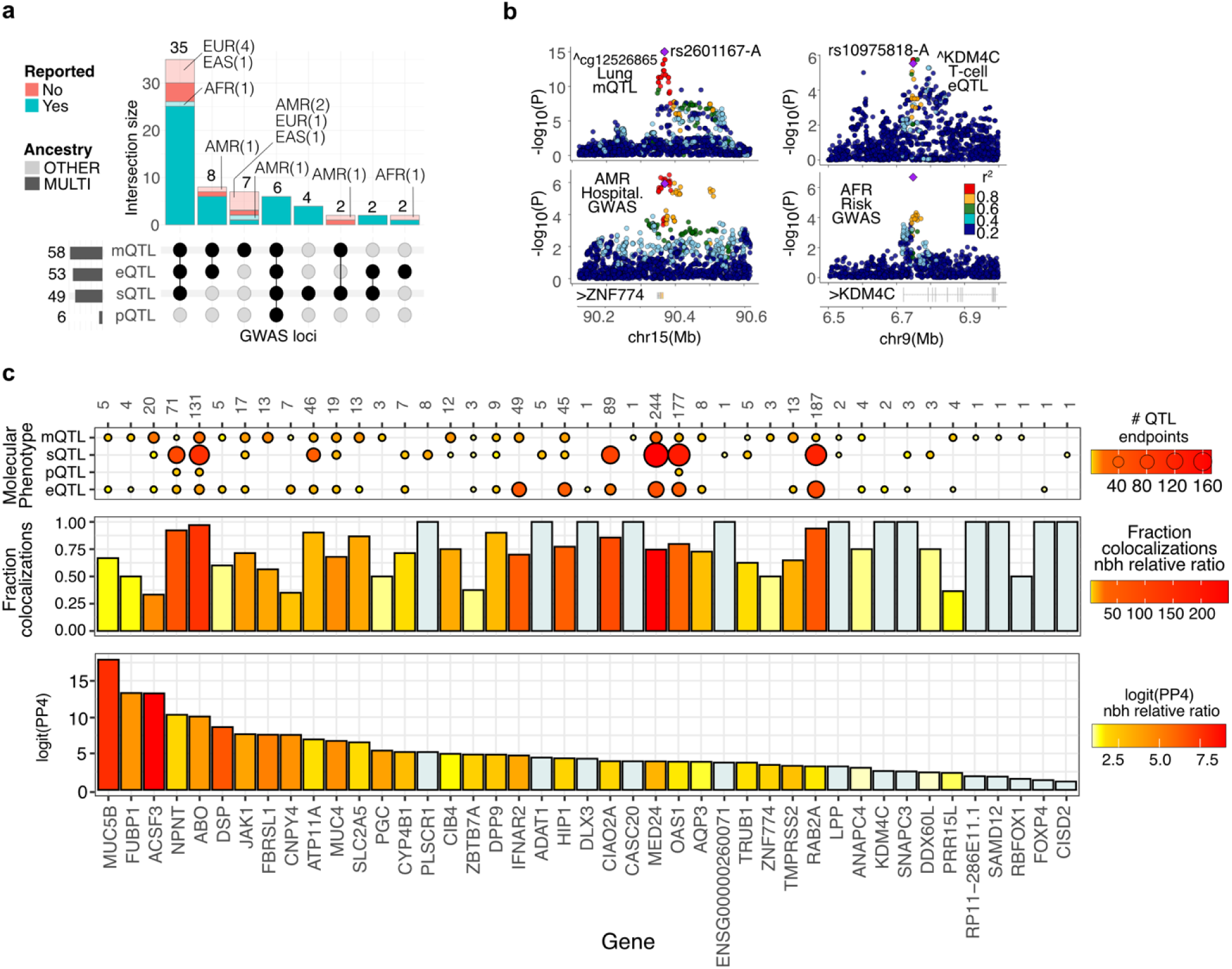
COVID-19 GWAS-QTL colocalization results. **a**, Number of COVID-19 GWAS loci with identified QTL colocalizations, stratified by QTL molecular phenotype, GWAS ancestry, and previously reported status. **b**, Genotype-phenotype association p-values of the ZNF774 and KDM4C locus. Left panels illustrate CpG cg12526855 mQTL signal in Lung (top) and GWAS signal for COVID-19 severity (Hospitalization GWAS) in the Admixed American cohort (bottom). Right panels illustrate KDM4C eQTL signal in T-cells (top) and GWAS signal for COVID-19 risk in African cohort (bottom). Top variant by posterior probability of shared causality is illustrated with a diamond and labelled by dbsnp id and COVID-19 risk/severity associated allele. ‘v’ and ‘L’ indicate allele association with negative or positive molecular phenotype effect, respectively. ‘>’ and ‘<’ indicate gene transcription direction. Linkage disequilibrium between loci is quantified by squared Pearson coefficient (r2), derived from corresponding GWAS ancestry. P-values correspond to nominal GWAS and QTL associations, derived from multiple regression two-sided t-tests. **c**, Colocalization evidence metrics (y axis) stratified by gene (x axis). Bottom panel illustrates the logit(PP4) corresponding to the top per-locus gene for that metric; bars are colored by the relative difference (ratio) with the corresponding locus second-ranked gene for the same metric. Middle panel illustrates the per-locus fraction of colocalizations attributable to corresponding gene; bars are colored by the relative difference (ratio) with the corresponding locus second-ranked gene for the same metric. For both panels, genes corresponding to loci with a single gene colocalized are colored in light blue. Top panel illustrates the number of QTL endpoints (QTL maps) per gene and molecular phenotype. Total number of colocalizations per gene is displayed on top of corresponding slot. Genes are sorted by logit(PP4) metric.

We identified between 1 and 19 COVID-19 colocalized genes per locus (median = 4) and observed that most (>81%) loci support colocalization for multiple genes, and that at half of these multi-gene loci, a single gene accounts for the majority of colocalizations. Moreover, most loci (86%) implicate colocalizations across multiple QTL cell/tissue endpoints (Supplementary Table 5, Supplementary Figure 1). These phenomena are expected, due to varying regulation between cell types and widespread coregulation ^45,46^, among other causes. However, it makes identification of putative causal gene(s) difficult, as most colocalizations are expected to not be causal ^47^. To prioritize candidate genes by colocalization evidence, we calculated per-gene metrics based on high posterior probability of colocalization logit(PP4) and on accounted fraction of colocalizations (Methods, Supplementary Table 5). We observed that the colocalization metrics agree only in part; while 67% of 64 gene-mappable colocalized loci have a gene candidate supported by both metrics, genes that rank high in one metric do not necessarily rank high in the other (Fig. 3c). For instance, *TMPRSS2* and *OAS1*, well-known COVID-19 linked genes, account for >60% of corresponding locus colocalizations, but have a relatively moderate probability of colocalization compared to other genes. Relative gene-prioritization score differences are employed to prioritize causal genes ^25,48^; here we calculated, per metric, the relative difference with the secondmost supported gene per locus. We observe that while the colocalization fraction scores are only marginally correlated (r=0.31, P=0.03), the relative colocalization probability scores tend to agree with the absolute ones (r=0.79 and P=2.2e-16), providing limited additional information for prioritization purposes.

The amount of colocalization signal attributable to a gene can reflect the shared impact of the underlying causal QTL variant across biological contexts, and is associated to higher chances of organismal-level phenotypic pleiotropy ^46^. Multiple regulatory effects (e.g. on splicing and expression) for the same gene often mediate the same complex trait associations, and QTLs derived from different molecular phenotypes can have an independent contribution to complex traits ^21,46^. Here we have quantified the amount of total and cross-omics colocalization support per gene to evaluate the agreement with the aforementioned colocalization evidence metrics, and to further aid putative causal gene prioritization. We observe that genes supported by both metrics tend to account for a higher number of colocalizations (Wilcoxon rank sum test P=1.7e- 07) and are more likely supported by more than one QTL molecular phenotype (Wilcoxon rank sum test P=4.45e-11), than other colocalized genes. The only three genes supported by all molecular phenotypes analyzed are *NPNT*, *ABO* and *OAS1* (Fig. 3c), all well-known COVID-19 linked genes. Altogether, we provide complementary colocalization evidence metrics to help prioritize putative causal COVID-19 genes.

To enable further exploration of the colocalization signals identified here, we built an interactive R shiny application (https://covidgenes.shinyapps.io/shiny/) to easily query summary statistics and visualize the genomic context of all significant QTL colocalizations, stratified by QTL map, molecular phenotype, and gene (Supplementary Figure 2a). Using this tool to contextualize colocalization signal, we identified genes with suboptimal colocalization evidence scores that are arguably likely causal candidates. For instance, in the rs8192330-G|BMP1 locus, *BMP1* and *SFTPC* account for 5/10 and 1/10 of colocalizations, respectively. The top – largest PP4 – colocalization instance corresponds to *SFTPC* differential splicing identified exclusively in lung (Supplementary Figure 2b-c). Other candidates in the region, which have lower colocalization probabilities, do not account for colocalization signal in lung and are not part of mechanisms known to play a role in COVID-19. Overall, the application allows locus-level contextualization of colocalizations that can assist the identification of putative causal genes.

### Pulmonary cell of origin of COVID-19 linked genes

SARS CoV 2 transmission and infection occurs via the respiratory system, and COVID-19 affects lung more than other organs ^49^. Thus, it is expected that a large fraction of causal molecular perturbations occurs in lungs. We observed that lung accounts for a relatively small fraction of colocalizations, and effect sizes of loci colocalizing in lung are not particularly large (Supplementary Figure 1). However, we identified lung as the most overrepresented biotype when comparing biotype-stratified colocalization signal with other GWAS trait endpoints from the Open Target database (Supplementary Figure 3a). To investigate the pulmonary cell of origin of COVID-19-gene associations, we adopted two complementary approaches, based on lung cell-type interaction *cis* eQTLs (ieQTLs) and lung single-cell RNA-Seq *cis* eQTLs (sc-eQTLs) (Methods). Cell-type ieQTLs are derived from bulk RNA-Seq profiles and capture eQTLs correlated with cell-type abundances ^50,51^; sc-eQTLs capture eQTLs in specific cell types identified from single-cell RNA-Seq profiles ^52^. Both approaches are complementary, as ieQTLs can misattribute signal to cell types correlated with the targeted one, and sc-eQTLs can miss signal corresponding to rarer cells and lowly expressed genes.

First, we integrated 18 COVID-19 bulk lung eQTL colocalized variant-gene pairs with ieQTL maps generated herein (Methods) from pulmonary epithelial, endothelial, stroma and macrophage cell scores ^51^. We observe enrichment for ieQTL signal (Fig. 4a), which suggests that the genetic regulation of COVID-19 linked genes in lung tend to occur in a cell-type specific manner. Epithelial cells are enriched the most, in agreement with being the principal target of SARS CoV 2 infection in the distal lung ^53^. We detect 10 significant (FDR<=0.10) ieQTLs corresponding to six genes (Fig. 4b), for which the COVID-19 pathological allele increases (rs383510:T-*TMPRSS2*, rs3859191:G-*GSDMA*, rs3848456:A-*DLX3*, rs60840586:A-*AQP3*) or decreases (rs35705950:G-*MUC5B*, rs2076295:G-*DSP*) transcript abundance. While multiple lines of evidence support a mechanistic link between *TPRSS2* and *MUC5B* and COVID-19, links with the other ieQTL genes have been less, or not, characterized. Among those, we identified *AQP3* and *DLX3*, with COVID-19 colocalized eQTLs specific to pulmonary epithelial cells (Fig. 4b, Supplementary Figure 3b). In skin, *DLX3* can upregulate proinflammatory cytokines and favor the accumulation of macrophages ^54^; it may play similar function in COVID-19 infected lung tissue. While a genetic COVID-19-lung-DLX3 link has been reported ^55^, specificity for pulmonary epithelial cells, described here, has not. In COVID-19-infected lung, aberrant proliferation of AQP3*+* epithelial basal cells has been reported ^56^, and aquaporins have been proposed as COVID-19 drug targets ^57^. Here, we describe a robust, multi-omic genetic AQP3- COVID-19 association (Fig. 3c) enhanced in pulmonary epithelial cells (Fig. 4b, Supplementary Figure 3b), pinpointing *AQP3* as a putative COVID-19 causal gene.

**Fig. 4.**
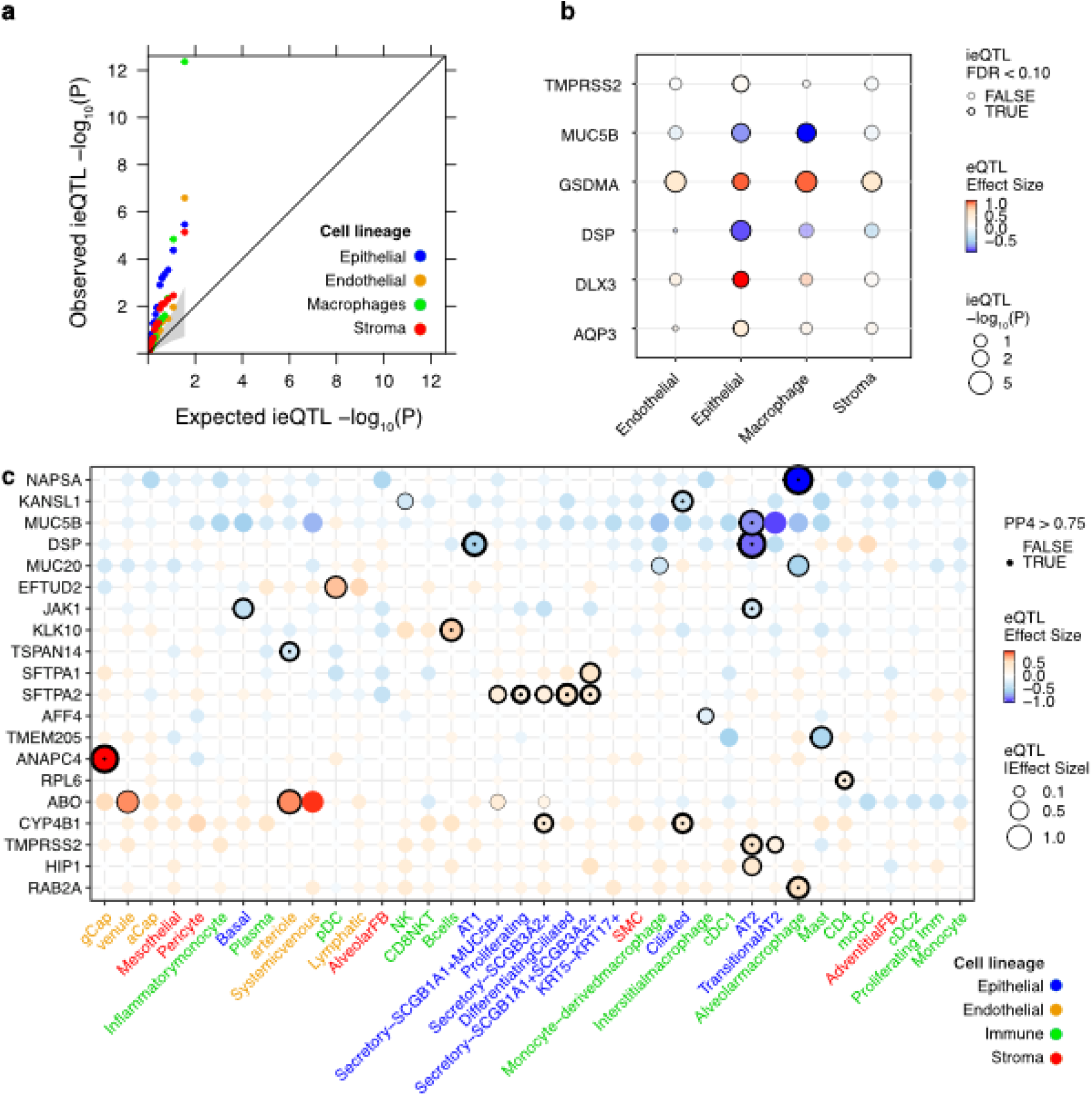
Pulmonary cell of origin of COVID-19 linked genes. **a,** Quantile-quantile plots for lung cell-type ieQTL p-values corresponding to genotype by cell-type abundance interaction term. Analysis was focused on 18 COVID-19 bulk lung eQTL colocalized variant-gene pairs. The shaded area indicates the 95% confidence interval expected under the null. **b,** QTL summary statistics for significant (FDR<0.10) ieQTL genes (x axis) by pulmonary cell type (y axis). The eQTL effect size was mapped from cell-type enriched samples and FDR was assessed at the level of SNP-gene pair (Methods). **c,** Colocalization scores and eQTL effect sizes for genes (y axis) corresponding to sc-eQTL-COVID-19-GWAS colocalizations (PP4>0.5) in at least one pulmonary cell type (x axis). Labels of cell types of epithelial, endothelial, immune, and stromal origin are typed in blue, orange, green and red, respectively. Gene-cell pairs tested for colocalization are indicated with stroked circles. Increased thickness of the stroke corresponds to higher PP4 values. Gene-cell pairs with strong (PP4>0.75) colocalization signal are indicated with a dot. Rows and columns were clustered by complete clustering based on eQTL effect size dissimilarity estimated by euclidean distance. In b-c, i/eQTL effect sizes correspond to COVID-19 pathological allele.

Next, we colocalized COVID-19 GWAS hits with sc-eQTLs maps from 38 lung cell types (Methods) and identified 28 suggestive (PP4>=0.50) colocalizations, 17 of which have strong (PP4>=0.75) colocalization support (Fig. 4c, Supplementary Table 5). Like ieQTL enrichments, most (17/28) colocalizations are identified in epithelial cells; eight and three colocalizations were identified in immune and endothelial cells, respectively. We identified genes and processes with strong evidence of a causal role in COVID-19, including *TMPRSS2*, related to viral entry, *MUC5B*, involved in muco-ciliary clearance of pathogens, *JAK1*, involved in cytokine release, and *SFTP1, SFTP2* and *NAPSA* genes, involved in surfactant metabolism. We observe marked cell subtype specificity; *SFTP* genes and *NAPSA* are markers of AT2 cells, but corresponding COVID-19 colocalizatoin were exclusively detected in secretory and alveolar macrophage cells, respectively. *ANAPC4* colocalization was found exclusively in general capillary (gCap) cells. These highly specialized endothelial cells act as progenitor cells to the endothelium, maintain capillary homeostasis, and regenerate damaged aerocytes in COVID-19-injured epithelium ^58^. The role of cell cycle gene *ANAPC4* in such process has not been characterized and warrants further investigation. Finally, we identified the uncharacterized COVID-19 severity rs2076295-G colocalization as linked to lower *DSP* expression in alveolar cells, exclusively. Importantly, the colocalization signal for these reported uncharacterized associations is very strong (PP4>=0.98) (Supplementary Figure 3c-e).

Together, the results presented pinpoint uncharacterized COVID-19 gene associations in lung and contribute to the understanding of the cell-type specificity of COVID-19 pulmonary pathobiology, including the mechanism of action of implicated genes.

## Discussion

In this work, we sought to identify human genes influencing genetically driven COVID-19 risk and severity, and to identify additional high-order phenotypes impacted by pleiotropic COVID- 19-associated genomic loci, with emphasis on characterizing the pulmonary pathobiology of the disease.

While comprehensive characterization of the shared genetics among COVID-19 and other complex diseases and traits, using different phenome-wide approaches, has provided insights into the pathobiology of COVID-19 ^11,59^, our analysis constitutes the most extensive effort to date of integrating COVID-19 with other trait GWAS signals, via a colocalization approach. In addition to improving basic understanding of COVID-19 biology and host interactions, this work may inform future therapeutic mechanisms for COVID-19 in two ways. First, by illuminating some of the detailed molecular mechanisms through which human genetics influences COVID- 19 risk and severity, we provide templates for therapeutic approaches that likely have the higher probability of success in clinical development (King et al. 2019, Minikel et al. 2024). Second, with comprehensive characterization of shared genetics between COVID-19 and other human traits, we might separate mechanisms expected to influence COVID-19 uniquely from those that might have unintended effects on other aspects of human biology.

While overlap of COVID-19 GWAS signal with pulmonary traits has been described ^11^, here we confirm their disproportionate representation, and evaluate per-trait per-locus consistency of directionality of effect with COVID-19 pathological alleles. Consistent detrimental or protective effects are observed for lung adenocarcinoma and COPD, respectively. The observed opposite allelic effects suggest the presence of genetic effect trade-offs in the lung, where inherited pulmonary physiological and molecular features that decrease likelihood to suffer from COPD increase chances to suffer from severe COVID-19. These phenomena are hypothesized to depend on cell type, lung area and age ^60^. For IPF and lung function metrics, observed locus-dependent detrimental or protective effects how COVID-19 pathology is connected to lung function and respiratory diseases by complex pleiotropic relationships.

This study comprises the most extensive effort of integrating molecular QTL sources with COVID-19 phenotypes to date. The usage of an extensive, multi-molecular-phenotype QTL catalog, including cell-type resolution maps derived from causal cell-of-origin contexts – e.g. pulmonary cells - maximizes the probability of detecting trait-causal genes and contexts at a cost of low specificity, i.e. detecting non-causal associations. To facilitate causal gene prioritization, we generated a resource (R shiny app) that provides a high-level view of all colocalization signal per locus, enabling context-aware evaluation of colocalization evidence. Additionally, we generated aggregated, complementary colocalization evidence metrics that we utilize to flag highly supported genes that may be most likely to be causal. Reassuringly, among high-scoring genes, we identified well-known COVID-19-linked genes. We also find candidate causal genes of uncharacterized loci, e.g. DSP. While QTLs are unequivocally useful to identify causal genes given GWAS signal, their predictive power is leveraged and complemented when combined with functional annotations ^25^, in part because GWAS hits may not have detectable QTLs ^20,61^. The colocalization evidence resources generated herein can be utilized for that purpose.

Selection of a reasonably liberal GWAS significance threshold ^62^ aimed to - at a cost of considering spurious associations - maximize the inclusion of loci that despite weak GWAS signal, when integrated with additional sources can point at molecular and phenotype links that contribute to the understanding of COVID-19 biological mechanisms. The desmoplakin- encoding DSP locus constitutes a notable example of such cases, given pulmonary trait links, high colocalization evidence and pulmonary cell associations observed. In combination with what is known about the shared ethiology of both diseases ^63^ and desmoplakin function ^26^, our results point to DSP-driven COVID-19 and IPF shared genetic perturbations, possibly acting on fibrogenesis and maintainance of airway epithelial integrity.

Altogether, we contributed to providing molecular and phenotype links to the genetic architecture of COVID-19 risk and severity phenotypes, by further characterizing previously reported loci and by providing novel insights for uncharacterized loci. Importantly, causal relationships between COVID-19 and genetically correlated phenotypes and molecules are pinpointed but not interrogated here; dedicated statistical approaches - such as mendelian randomization – and functional characterization could be applied for that purpose.

## Methods

### Determination of GWAS significant loci of COVID-19 disease risk and severity

To enable the characterization of the genetic mechanisms underlying COVID-19 risk and severity on human host, we used GWAS summary statistics generated by the COVID-19 Host Genetics Initiative Consortium (HGI) ^23^. The COVID-19 GWASs correspond to worldwide meta-analyses, without 23andMe, version 7, released on April 8, 2022; data production and quality control are described in detail in ^11^. Three COVID-19 GWAS phenotypes were considered, as defined by HGI: Critical illness (A2), comparing confirmed COVID-19 infected individuals experiencing very severe respiratory symptoms vs. population, Hospitalization (B2), comparing hospitalized individuals due to COVID-19 infection to population, and Risk (C2), comparing confirmed COVID-19 affected individuals vs. population. We assume A2 and B2 GWASs to be broadly informative about genetic determinants of COVID-19 severity, and C2 to better capture the genetic susceptibility to SARS-CoV-2 infection. The GWAS summary statistics from each COVID-19 GWAS phenotype are derived from six ancestry endpoints that include one multi-ancestry, and five ancestry-stratified cohorts (South Asian (SAS), East Asian (EAS), African (AFR), Admixed American (AMR), European (EUR)). Of note, COVID-19 HGI GWAS B1, comparing hospitalized vs. non-hospitalized individuals due to COVID-19 infection, was not considered, as summary statistics were only available for the multi-ancestry but not for the ancestry-stratified GWAS approach. In total, 18 (3 COVID-19 GWAS phenotypes for 6 ancestry endpoints) were considered; number of cases and controls are detailed below.

| Phenotype | Multi-ancestry | EUR | AMR | AFR | EAS | SAS |
| --- | --- | --- | --- | --- | --- | --- |
| **Critical illness (A2): confirmed COVID-19 infected individuals experiencing very severe respiratory symptoms vs. population** | 18,152 vs. 1,145,546 | 13,769 vs. 1,072,442 | 769 vs. 6209 | 628 vs. 12,568 | 794 vs. 4,862 | 980 vs. 47,696 |
| **Hospitalization (B2): hospitalized individuals due to COVID-19 infection vs. population** | 44,986 vs. 2,356,386 | 32,519 vs. 2062805 | 3,077 vs. 66,686 | 2589 vs. 123,225 | 2,882 vs. 31,200 | 1,622 vs. 47612 |
| **Risk (C2): confirmed SARS-CoV-2 infected individuals vs. population** | 159,840 vs. 2,782,977 | 122,616 vs. 2,475,240 | 10,377 vs. 74556 | 8,814 vs. 129140 | 4,459 vs. 36,121 | 8,945 vs. 44,171 |

We identified autosomal loci with suggestive GWAS signal, i.e. genomic windows containing GWAS signals, across phenotypes and ancestry endpoints. We first constructed a reference dataset of best-guess genotypes from UK Biobank (UKBB)^64^ by considering imputed dosages of variants with info score > 0.3 and MAF > 0.1%, selecting genotypic data corresponding to 15,000 randomly selected or to 2,000 ancestry-matched unrelated UKB samples, to generate multi-ancestry or single-ancestry genotype panels, respectively. We filtered variants with missingness > 5% and Hardy–Weinberg equilibrium test P < 1 × 10−7. For each of the 18 GWASs, we used the PLINK ref ‘clumping’ algorithm to select top-associated variants (P < 5 × 10−7) and corresponding LD-linked variants at r2 > 0.05 with the top associated variant within ±1 Mb, utilizing the GWAS-matching ancestry-stratified or multi-ancestry UKB genotype. We determined the genomic span of each LD-based clump and added 1 kb up- and downstream as buffer to the region. If any of these windows overlapped, we merged them together into a single (larger) locus.

To determine a set of non-redundant GWAS loci across ancestries and COVID-19 phenotypes, we first merged overlapping clumps per ancestry across phenotype endpoints (A2, B2, C2). Then, we selected all multi-ancestry derived clumps, and complemented this set with non-overlapping clumps identified in a single ancestry. The resulting set of loci is composed of genomic regions with suggestive GWAS signal in at least one phenotype endpoint in at least one ancestry endpoint. For each GWAS locus, considering the ancestry endpoint where the GWAS hit was defined, we selected a ‘top’ GWAS across phenotype endpoints as the GWAS with the smallest p-value per locus lead variant, which we define as the top GWAS lead variant. Next, we selected GWAS hit loci with at least four variants with top GWAS P<1-04 and MAF>0.01. Of note, this filtering excludes the MUC16 locus, as it does only have three variants passing the filter. Finally, we filtered out GWAS hit loci overlapping the Major Histocompatibility region (MHC); the final set is composed of 91 COVID-19 GWAS loci. Unless stated otherwise, for each GWAS locus, the summary statistics of top GWAS was utilized in subsequent analyses. Top GWAS loci lead variants were annotated with the nearest (closest transcription start site from canonical transcript) protein-coding gene, with VEP v.111 ^24^ predicted impacted gene and corresponding most severe impact type, and with variant-to-gene (V2G) ^25^ predicted impacted gene, i.e. corresponding to top V2G score. V2G predictions and nearest protein-coding gene annotation were obtained from Open Targets Genetics v.8 ^65^.

### Overlap with previously reported COVID-19 GWAS loci

We sought to determine which of the 91 COVID-19 GWAS loci analyzed here overlap with previously reported COVID-19 GWAS loci. First, we obtained EBI GWAS Catalog entries (https://www.ebi.ac.uk/gwas; 2024-04-05) at GWAS P <5e-7 that matched COVID-19 phenotypes (MONDO:0100318 and EFO:0600019 descendants, EFO v.3.64.0). We obtained 14,102 entries corresponding to 39 studies and 156 GWAS endpoints. Additionally, since they were not present in the catalog, we added entries from ^11^: COVID-19 GWAS loci that the COVID-19 HGI Consortium identified at P<5e-08 from the version 7 data release, which correspond to the multi-ancestry GWAS summary statistics endpoints utilized herein. For each of the 91 COVID-19 GWAS loci, we intersected the region analyzed in this paper with this set of previously reported COVID-19 entries to determine ‘Reported’ status.

### Determination of the severity versus risk score

To identify which genomic loci were more likely to be associated with SARS-CoV-2 infection susceptibility or with COVID-19 severity, we employed the same approach utilized by the COVID-19 Host Genetics Initiative: a two-class Bayesian model based on the patterns of association across two COVID-19 GWAS phenotypes, B2 (severity) and C2 (infection risk). This approach calculates a posterior probability for each proxy GWAS lead variant to more likely be associated with infection risk or severity. We classified variants as severity- or infection-associated if their score was less than 0.2 or its inverse - greater than 0.8, respectively; scores between 0.2 and 0.8 are considered ambiguous. This method is described in ^12^, corresponding code is hosted in https://github.com/mjpirinen/covid19-hgi_subtypes.

### Colocalization of COVID-19 GWAS with human trait GWAS maps

To investigate the relationship between COVID-19 associated loci and organism-level human traits, we used *POEMcoloc* ^37^, a method for colocalization analysis with incomplete summary statistics. We downloaded the complete GWAS catalog in February 2023. To apply *POEMcoloc*, we first derived from information in the GWAS catalog, broad ancestry group category, GWAS design (case/control vs quantitative), sample sizes, and case fraction for each of 487,213 entries. In order to be tested, a GWAS catalog entry was required to have all necessary elements for *POEMColoc* and to be genome-wide significant (P<5E-8). Most entries excluded from analysis were due to the significance threshold rather than missing information and in total, 353,869 GWAS entries were candidates for colocalization, we refer to them as ‘GWAS loci’.

For each of the 91 COVID-19 associated loci, we used summary statistics from the top GWAS, and the information extracted for each overlapping GWAS catalog locus, to determine colocalization probabilities with *POEMColoc.* Of 91 COVID-19 loci, two had no overlapping GWAS catalog loci, while the remaining loci had between 1 and 1316 overlapping loci. There were a total of 13,833 testable pairs of COVID-19 associated locus/GWAS catalog locus, of which 13,750 corresponded to non-COVID-19 phenotypes and were used for subsequent analyses. We imputed summary statistics for each GWAS catalog locus with *POEMColoc* and LD information derived from either all unrelated individuals in the 1000 genomes reference panel, or the appropriate population-specific sub-panel in cases where the GWAS catalog locus could be confidently assigned to a specific ancestry group. We defined suggestive support for colocalization between the COVID-19 and other trait GWAS signal at posterior probability for a single shared causal variant PP4>0.75. Colocalizations corresponding to GWAS Catalog COVID-19 phenotypes (MONDO:0100318, EFO:0600019 from EFO v. 3.51.0, and corresponding descendants) were excluded.

### Annotation of GWAS trait endpoints

Mappings from GWAS trait endpoints to EFO were obtained directly from the GWAS Catalog. When a trait was mapped to multiple EFO terms, we used their common ancestor. Using the EFO ontology (EFO v.3.51.0) and the *ontologyIndex* R package v. 2.10.0 [https://cran.r-project.org/web/packages/ontologyIndex/citation.html] we mapped each association to EFO ancestor terms as well as the originally reported EFO trait. Mapping to ancestral terms allows us to look at colocalization frequency for broader categories of traits, for example, “lung diseases.” To generate Figure 2a, GWAS Catalog EFO terms were assigned to the higher-level category. Some categories encompass multiple EFO ancestors: cardiovascular and lipid traits include ’cardiovascular disease’, ’cardiovascular measurement’, ’blood pressure’, ’lipid or lipoprotein measurement’; digestive system disease and measurement includes ’digestive system disease’, ’liver enzyme measurement’, ’abnormality of the digestive system’; nervous system and cognition includes “brain measurement”, “neuroimaging measurement”, “cognition”, “cognitive function measurement”, ’educational attainment’, ’nervous system disease’, ’mental or behavioural disorder biomarker’, ’pain’; and respiratory diseases and measurements includes “respiratory system disease”, ’respiratory disease biomarker’, ’pulmonary function measurement’. If multiple categories could have been assigned, the category corresponding to the most frequently colocalized ancestor was used. In Figure 2b, we consider non-redundant EFO terms with four or more colocalizations. EFO terms are considered redundant if they are ancestors of reported EFO traits corresponding to the same set of clumps as another, more specific descendant EFO term. For example, macular degeneration was considered redundant with the more specific term age-related macular degeneration because all macular degeneration associations were also found for age-related macular degeneration. Association between per-loci pleiotropy, measured by number of colocalized GWAS endpoints, and severity versus risk score, was calculated by Spearman’s rank correlation implemented in *cor.test* R v.4.1.2 function.

### Imputation of direction of effect at GWAS loci

We annotated GWAS Catalog associations with direction of association between the COVID-19 reported allele and the GWAS Catalog trait, where possible. For diseases, the risk allele corresponds to increased disease risk. For quantitative traits, we used the field “95% CI (TEXT)” to determine whether the risk allele was associated with increased or decreased trait values. In cases where the risk variant reported in the GWAS catalog was the same as the COVID-19 risk variant, we checked for a match between the reported alleles. Otherwise, we computed the correlation between the two risk alleles in the 1000 genomes phase 3 reference panel to determine if they were positively or negatively associated. We used the 1000 genomes superpopulation corresponding to the reported ancestry from the GWAS Catalog, and if multiple broad ancestral categories, 1000 genomes unrelated individuals from all ancestry groups. Combining this with the reported direction in the GWAS Catalog, we annotated the direction of association between the COVID-19 risk allele and the GWAS trait.

### Determination of molecular QTL-COVID-19-GWAS colocalized loci

To investigate possible associations between cis-genetically regulated molecular phenotypes (QTLs) and COVID-19, we compiled an exhaustive QTL map collection and employed different colocalization approaches.

### Compilation of molecular QTL full-summary statistics maps

To maximize the expectation of identifying COVID-19 putatively causal molecular links, we compiled an exhaustive collection of *cis* quantitative trait loci (QTL) mappings (maps) derived from several molecular phenotypes (MPs): gene (eQTLs), splicing phenotypes (sQTLs), DNA methylation (mQTLs) and protein abundance (pQTLs).

The QTLs originate from widely different contexts, i.e., tissue and cell types, stimulus and developmental states; we considered a total of 322 *cis* QTL maps with full statistics available and genome-wide molecular phenotype tests. The majority (94%) of QTL maps are derived from gene expression or splice phenotypes (e/sQTLs), 4% are derived from DNA methylation (mQTLs) and 2% from protein (pQTLs) abundances. Details of QTL maps are provided in Methods, Supplementary Table 3. A total of 158 eQTL maps were obtained from bulk-tissue or isolated cells, 127 of which from 31 different studies included in the eQTL Catalogue (https://www.ebi.ac.uk/eqtl/, version 5, April 2022. A total of 31 additional bulk-tissue and isolated-cell eQTL maps were obtained from four additional sources, derived from whole blood from individuals unaffected by COVID-19 (eQTLGen, https://eqtlgen.org/cis-eqtls.html) as well as from COVID-19 patients (https://humandbs.biosciencedbc.jp/en/hum0343-v2#qtl), induced pluripotent stem cells (iPSC) (i2QTL, https://doi.org/10.5281/zenodo.4005576) and 28 maps from isolated immune cells (ImmuNexUT, https://humandbs.biosciencedbc.jp/en/hum0214-v8#E-GEAD-420). In addition, lung eQTL maps derived from single cell RNA-Seq (sc-eQTLs) for 38 lung cell types, generated in ^66^, were obtained from GEO (GSE227136, mashr files). Considering splicing phenotypes, we included 109 sQTL maps derived from leafcutter phenotypes included in the eQTL Catalogue (https://www.ebi.ac.uk/eqtl/, version 6, April 2023). Considering DNA methylation, a total of 11 mQTL maps were obtained. We included 9 maps from eGTEx sources: breast mammary tissue, colon transverse, kidney cortex, lung, muscle skeletal, ovary, prostate, testis and whole blood (eGTEx, https://gtexportal.org/home/downloads/egtex), one additional muscle skeletal (FUSION, https://www.ebi.ac.uk/birney-srv/FUSION/) and one brain (ROSMAP, (http://mostafavilab.stat.ubc.ca/xQTLServe/) cis mQTL maps. Considering protein abundance, we included six pQTL maps from plasma (SomaScan deCODE 2021, https://download.decode.is/form/folder/proteomics, SomaScan Sun et al. 2018, https://www.ebi.ac.uk/gwas/downloads/summary-statistics; SomaScan and O-link FinnGen https://www.finngen.fi/en/access_results; ARIC EUR and AFR SomaScan http://nilanjanchatterjeelab.org/pwas).

### Colocalization of COVID-19 GWAS loci with molecular QTLs

For each of the 91 COVID-19 associated loci, we identified overlapping (> 1 bp) molecular phenotype (MP) *cis*-region loci from each QTL map. For each overlapping MP-GWAS region pair, we applied *coloc* v5.1.0.1 using molecular QTL and GWAS summary statistics as input. Analysis was performed only if the locus contained >=1 variant with nominal QTL P < 1e-05 and GWAS P < 5e-07. Prior probabilities of a variant yielding a) a QTL association (p1), b) a GWAS association (p2) and c) a QTL and a GWAS association (p12) were set to p1 = 1e-04, *p*2 = 1e-04, *p*12 = 1e-06. Only the regions with at least 50 variants in common between the GWAS and MP loci were tested for colocalization. Both for QTLs and GWAS statistics, colocalization was performed on effect size (effect size) and associated standard error (effect size s.e.) values. We defined suggestive support for QTL-COVID-19-GWAS colocalization at posterior probability PP4 >= 0.75. For mQTLs, CpG probe identifiers were mapped to genes according to regulatory region annotations from EPIC.hg38.manifest.tsv.gz and HM450.hg38.manifest.tsv.gz from https://zwdzwd.github.io/InfiniumAnnotation.

### Calculation of QTL-COVID-19-GWAS colocalization evidence metrics

We calculated per-gene metrics based on high posterior probability of colocalization and identified per-locus top gene for the metric. That is, we calculated the logit of PP4, and selected the top estimate per gene across corresponding colocalization instances. Additionally, for the top - largest logit(PP4) - gene per locus, we calculated the ratio between the gene metric and the metric corresponding to the next ranked gene. To measure the amount of colocalization signal attributable to a gene, we calculated the per-locus fraction of QTL maps implicated in the colocalizations derived from that gene. Additionally, for the top - largest fraction of colocalizations - gene per locus, we calculated the ratio between the gene metric and the metric corresponding to the next ranked gene. Entries for anti-sense genes were lumped together with complementary gene.

### Identification of biotype enrichment in eQTL-COVID-19-GWAS colocalized loci

To investigate whether COVID-19 GWAS-eQTL associations were enriched for a particular biotype class, we analyzed Open Targets ^25^, considering significant (P<0.75) GWAS-eQTL colocalizations. We considered eQTL colocalizations derived from 126 eQTL maps, mapped to 12 biotype classes, present both in our analysis and in Open Targets, as of December 2023 (Supplementary Table 3). We calculated the fraction of Open Targets GWAS phenotypes with a percent of eQTL biotype-associated GWAS loci larger than corresponding COVID-19 percent of eQTL biotype-associated loci and identified lung as a highly represented (top percent) biotype.

### Mapping of cell-type interaction eQTLs (ieQTLs) in lung

We mapped cell-lineage interaction *cis* eQTLs (ieQTLs) from N=515 GTEx v8 subject-matched bulk lung RNA-Seq profiles and genotype data ^46^ and cell-type enrichment scores’ for epithelial, endothelial, macrophage and stroma cells ^51^. Cell-type interaction eQTLs were mapped by fitting a linear regression model with an interaction term accounting for interactions between genotype and cell type enrichment scores:

p ∼ g + i + g ◦i + **C**

where p is the gene expression vector, g is the genotype vector, i is the inverse normal transformed xCell enrichment scores, and the interaction term g ◦ i corresponds to point-wise multiplication of genotypes and cell type enrichment scores. **C** is a matrix of covariates that were used for cis-eQTL mapping in lung ^46^ that include 60 PEER factors derived from gene expression, five genotype principal components, two covariates derived from the generation of genotype data by whole-genome sequencing (WGS) and biological sex status. The WGS covariates represent the WGS sequencing platform (HiSeq 2000 or HiSeq X) and WGS library construction protocol (PCR based or PCR-free). The filtered and normalized gene expression matrix corresponds to the one used for cis-eQTL mapping in lung ^46^. Briefly, gene expression values were inverse normal transformed after applying TMM for between-sample normalization. Interaction QTLs were identified by testing for the significance of the interaction term g ◦ i, and mapping was performed using an adaptation of v.2.184 FastQTL ^67^, available at https://github.com/broadinstitute/gtex-pipeline/tree/master/qtl. We restricted ieQTL mapping to 18 COVID-19 bulk lung eQTL colocalized variant-gene pairs, corresponding to 16 COVID-19 loci and 18 genes. To estimate cell-lineage eQTL effect sizes displayed in Fig. 4b, we mapped eQTLs by fitting p ∼ g + **C** on data derived from cell-type enriched samples, i.e., corresponding to the top tertile of xCell scores for each of the four ieQTL cell types, and with subject-matched genotype data. The ieQTL and eQTL effects were estimated considering the top colocalized variant per gene from COVID-19 bulk lung eQTL colocalizations.

### Colocalization of COVID-19 GWAS loci with lung sc-eQTLs

Bayesian colocalization analysis was performed using *coloc* v5.1.0.1 on COVID-19 GWAS hits and cross-cell meta-analyzed sc-eQTL summary statistics for each of the 38 lung sc-eQTL maps ^66^. Only protein coding genes were considered. For each overlapping MP-GWAS region pair, we applied *coloc* to sc-QTL along with GWAS summary statistics, only if the locus contained >=1 variant with nominal QTL P < 1e-04 and GWAS P < 5e-07. For GWAS statistics, colocalization was performed on effect size (effect size) and associated standard error (effect size s.e.) values. For sc-eQTL statistics, colocalization was performed on local false sign rate (LFSR) and MAF values. We defined suggestive and strong support for sc-QTL-COVID-19-GWAS colocalization at posterior probability PP4 > 0.50 and PP4 > 0.75, respectively.

## Availability of data and materials

The data sets analyzed during the current study are available from the following repositories: summary statistics of the COVID-19 GWASs (round 7) by the COVID-19 Host Genetics Initiative are available at https://www.covid19hg.org/results/r7/. The QTL summary statistics were obtained from multiple sources (see Methods). The variant calls from 1000 Genomes Project on the GRCh38 reference assembly are available from the UK Biobank Research Analysis Platform, accessible upon application approval. The summary statistics from the GWAS catalog are available at https://www.ebi.ac.uk/gwas/. Analysis and figures code, and figures data, has been deposited to https://github.com/AbbVie-ComputationalGenomics/COVID19_coloc_paper. R shiny is provided at https://covidgenes.shinyapps.io/shiny/ and https://github.com/AbbVie-ComputationalGenomics/COVID19_coloc_shiny

## Data Availability

https://github.com/AbbVie-ComputationalGenomics/COVID19_coloc_shiny

https://github.com/AbbVie-ComputationalGenomics/COVID19_coloc_paper

## Acknowledgements

We thank Jan Freudenberg, Mark Reppell, Mengzhen Liu, Justin E Ideozu, Vasanthakumar, Aparna Aparna Vasanthakumar, Sabah Kadri for critical review of the manuscript. We thank Howard J Jacob and IMI-CARE consortium for their feedback on corresponding manuscript analyses.

---

**Sup. Fig. 1.**
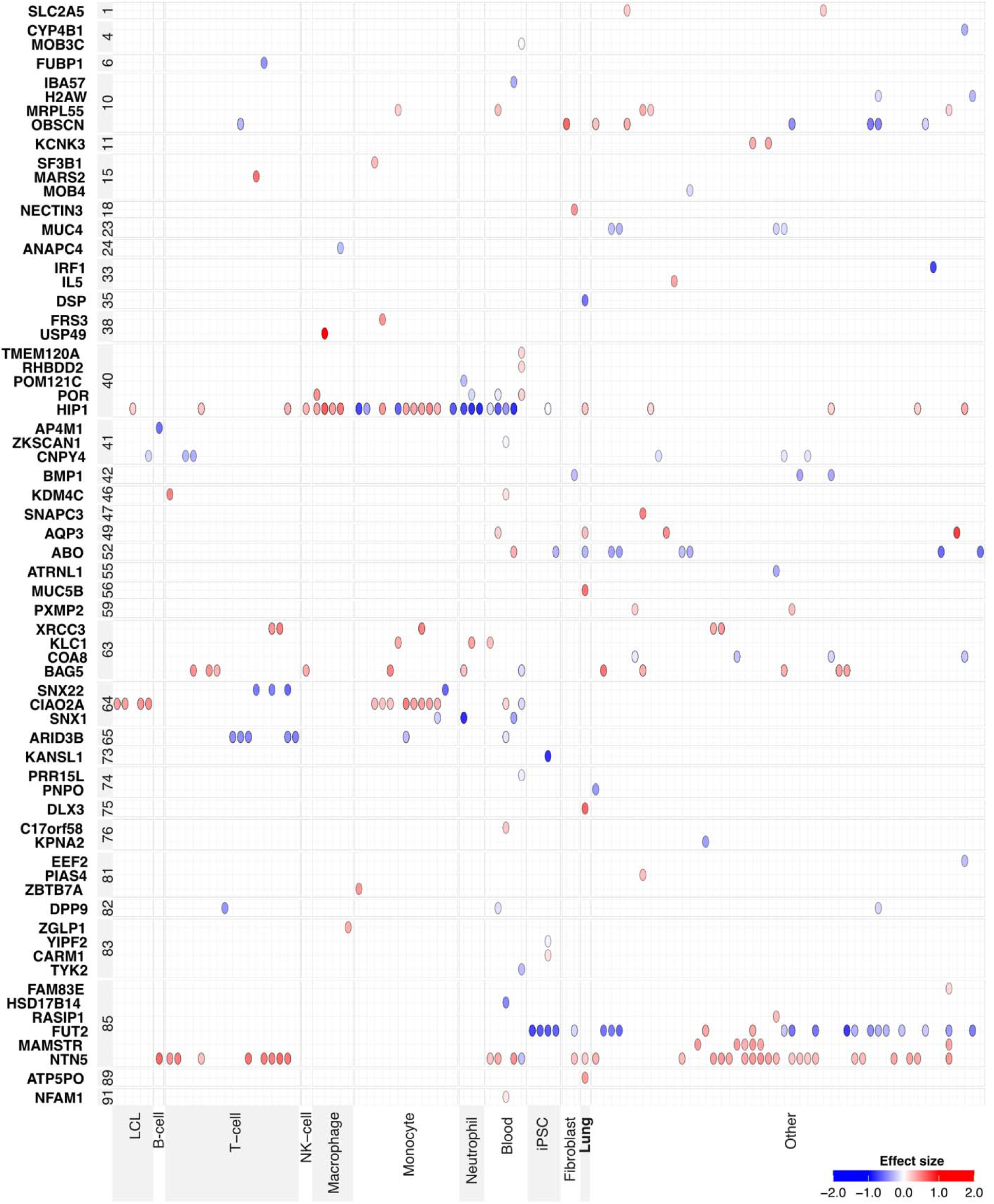
Biotype specificity of COVID-19 GWAS-QTL colocalizations. For significant (PP4>0.75) eQTL- COVID-19 GWAS colocalizations, the QTL effect size per colocalized protein-coding gene (y axis) and QTL map (x axis) is displayed. QTL effect sizes are mapped to the COVID-19 pathological allele and colored in red and blue for positive and negative effects, respectively; genes are grouped by corresponding GWAS locus index (Details provided in Supplementary Table 1). QTL maps are grouped and labelled by corresponding biotype category (Details provided in Supplementary Table 3).

**Sup. Fig. 2.**
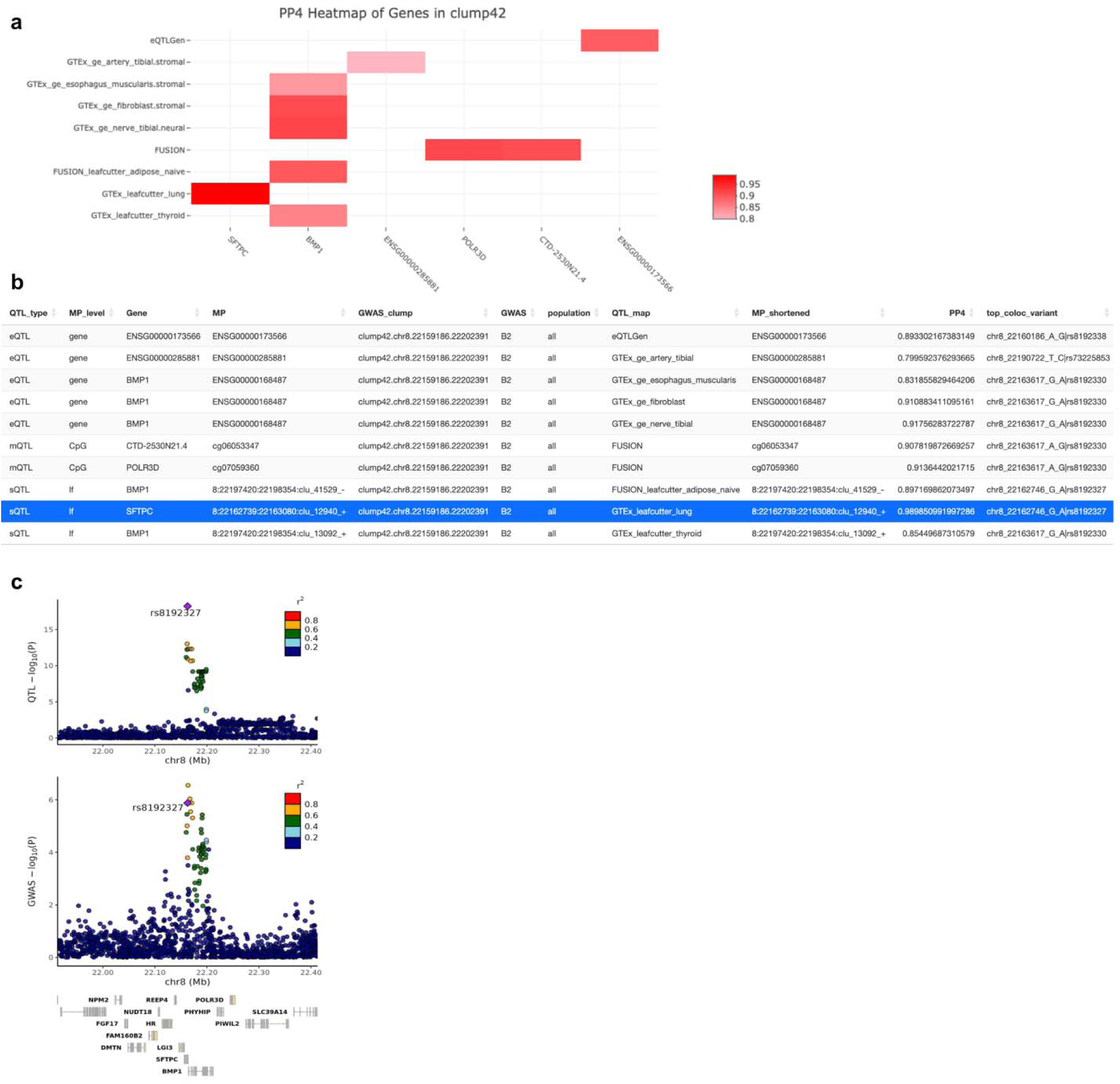
Visualization and overview of COVID-19-GWAS-QTL colocalization signal per locus. **a**, Figure panels adapted from the provided R shiny app interface https://covidgenes.shinyapps.io/shiny/. **a**, Significant COVID-19 GWAS-QTL colocalization instances (PP4>0.75) corresponding to the index number 42 COVID-19 GWAS locus, i.e. BMP1|SFTPC locus (Details in Supplementary Table 1), stratified by QTL map (y axis) and gene (x axis). **b,** COVID-19 GWAS-QTL features and summary statistics for *SFTPC* locus; *SFTPC* colocalization instance is highlighted. **c**, Genotype-phenotype association *P* values of the *SFTPC* locus. Panels illustrate *SFTPC* sQTL signal in lung (top) for the 8:22162739-22163080 *SFTPC* intron usage phenotype quantified by *LeafCutter*, and GWAS signal for COVID-19 B2 GWAS (bottom). Top GWAS-colocalized sQTL variant is typed in bold, linkage disequilibrium between loci is quantified by squared Pearson coefficient of correlation (*r*^2^), and colocalization probability (PP4) of sQTL with GWAS signal is shown. The diamond-shaped point represents the top (highest PP4) colocalized variant. P values correspond to nominal GWAS and sQTL associations, derived from multiple regression two-sided *t*-tests.

**Sup. Fig. 3.**
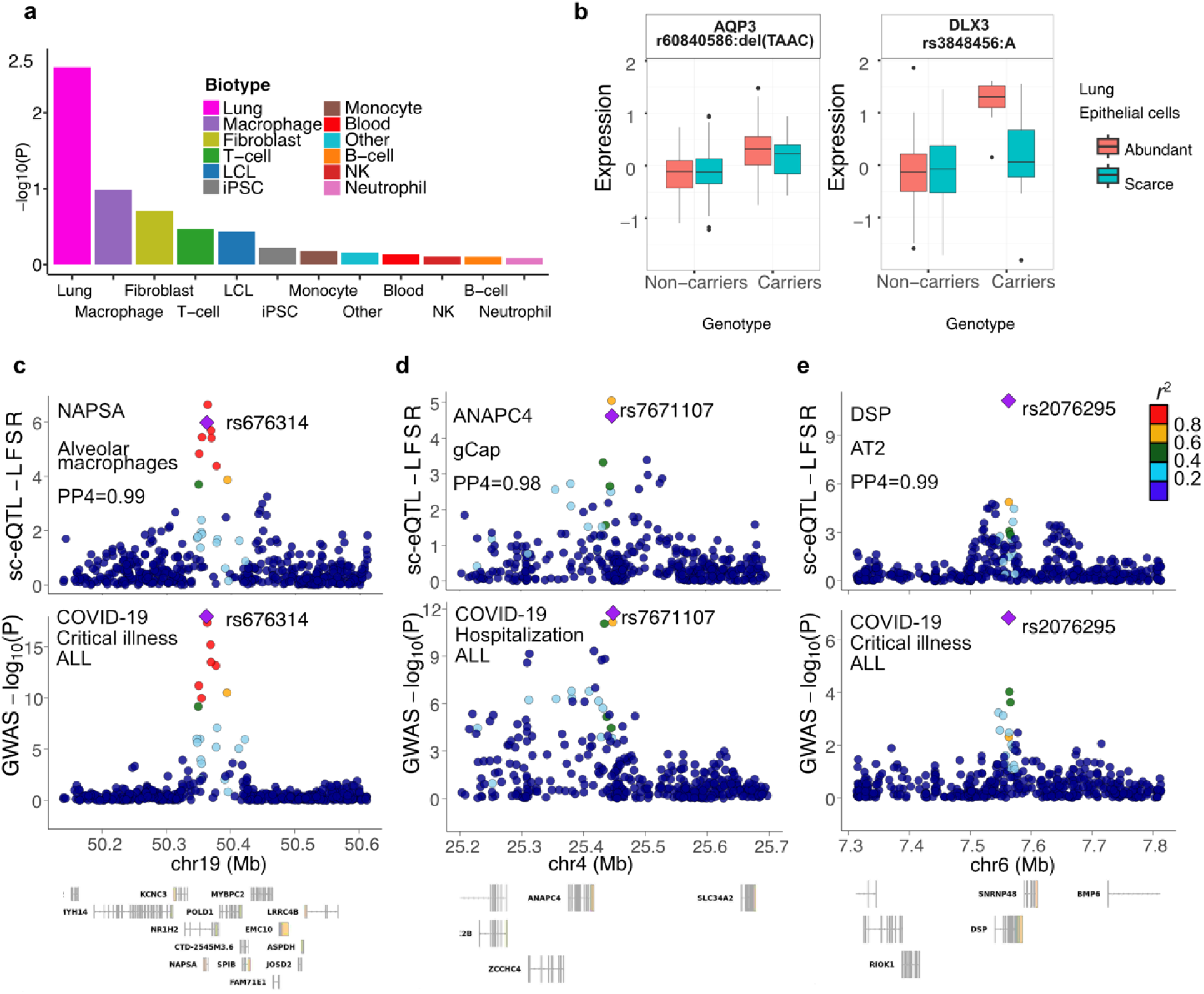
Pulmonary cell of origin of COVID-19 linked genes. **a)** Enrichment (y axis) of COVID-19 loci in eQTL biotypes (x axis). ‘P’ corresponds to the fraction of Open Targets GWAS phenotypes with a percent of eQTL biotype-associated GWAS loci larger than corresponding COVID-19 percent of eQTL biotype-associated loci (Methods). Gene **b)** Gene expression (y axis) of *AQP3* and *DLX3* genes by carrier status of corresponding COVID- 19 severity allele (x axis). Expression corresponds to GTEx v8 bulk lung RNA-Seq samples enriched (N=181) or depleted (N=156) in pulmonary epithelial cells, residualized by QTL mapping covariates (Methods). **c)** Genotype-phenotype association values of the *NAPSA* locus. Panels illustrate *NAPSA* sc-eQTL signal in pulmonary alveolar macrophages (top) and GWAS signal for COVID-19 critical illness GWAS (bottom). **d)** Genotype-phenotype association values of the *ANAPC4* locus. Panels illustrate *ANAPC4* sc-eQTL signal in pulmonary general capillary endothelial cells (top) and GWAS signal for COVID-19 Hospitalization GWAS (bottom). **e)** Genotype-phenotype association values of the *DSP* locus. Panels illustrate *DSP* sc-eQTL signal in pulmonary alveolar type II cells (top) and GWAS signal for COVID-19 critical illness GWAS (bottom). For panels c-e, linkage disequilibrium between loci is quantified by squared Pearson coefficient of correlation (*r*^2^), and colocalization probability (PP4) of sc-eQTL with GWAS signal is shown. Top variant (largest *coloc.abf* SNP.PP.H4) of GWAS-colocalized sc-eQTL is typed in bold and depicted by a diamond-shaped point. GWAS P values correspond to nominal GWAS, derived from multiple regression two-sided t-tests. LFSR correspond to sc-eQTL mashr local false sign rate values.

